# REACT-1 study round 14: High and increasing prevalence of SARS-CoV-2 infection among school-aged children during September 2021 and vaccine effectiveness against infection in England

**DOI:** 10.1101/2021.10.14.21264965

**Authors:** Marc Chadeau-Hyam, Haowei Wang, Oliver Eales, David Haw, Barbara Bodinier, Matthew Whitaker, Caroline E. Walters, Kylie E. C. Ainslie, Christina Atchison, Claudio Fronterre, Peter J. Diggle, Andrew J. Page, Alexander J. Trotter, The COVID-19 Genomics UK (COG-UK) Consortium, Deborah Ashby, Wendy Barclay, Graham Taylor, Graham Cooke, Helen Ward, Ara Darzi, Steven Riley, Christl A. Donnelly, Paul Elliott

## Abstract

**Background:** England experienced a third wave of the COVID-19 epidemic from end May 2021 coinciding with the rapid spread of Delta variant. Since then, the population eligible for vaccination against COVID-19 has been extended to include all 12-15-year-olds, and a booster programme has been initiated among adults aged 50 years and over, health care and care home workers, and immunocompromised people. Meanwhile, schoolchildren have returned to school often with few COVID-19-related precautions in place.

**Methods:** In the REal-time Assessment of Community Transmission-1 (REACT-1) study, throat and nose swabs were sent to non-overlapping random samples of the population aged 5 years and over in England. We analysed prevalence of SARS-CoV-2 using reverse transcription-polymerase chain reaction (RT-PCR) swab-positivity data from REACT-1 round 14 (between 9 and 27 September 2021). We combined results for round 14 with round 13 (between 24 June and 12 July 2021) and estimated vaccine effectiveness and prevalence of swab-positivity among double-vaccinated individuals. Unlike all previous rounds, in round 14, we switched from dry swabs transported by courier on a cold chain to wet swabs using saline. Also, at random, 50% of swabs (not chilled until they reached the depot) were transported by courier and 50% were sent through the priority COVID-19 postal service.

**Results:** We observed stable or rising prevalence (with an R of 1.03 (0.94, 1.14) overall) during round 14 with a weighted prevalence of 0.83% (0.76%, 0.89%). The highest weighted prevalence was found in children aged 5 to 12 years at 2.32% (1.96%, 2.73%) and 13 to 17 years at 2.55% (2.11%, 3.08%). All positive virus samples analysed correspond to the Delta variant or sub-lineages of Delta with one instance of the E484K escape mutation detected. The epidemic was growing in those aged 17 years and under with an R of 1.18 (1.03, 1.34), but decreasing in those aged 18 to 54 years with an R of 0.81 (0.68, 0.97). For all participants and all vaccines combined, at ages 18 to 64 years, vaccine effectiveness against infection (rounds 13 and 14 combined) was estimated to be 62.8% (49.3%, 72.7%) after two doses compared to unvaccinated people when adjusted for round, age, sex, index of multiple deprivation, region and ethnicity; the adjusted estimate was 44.8% (22.5%, 60.7%) for AstraZeneca and 71.3% (56.6%, 81.0%) for Pfizer-BioNTech, and for all vaccines combined it was 66.4% (49.6%, 77.6%) against symptomatic infection (one or more of 26 surveyed symptoms in month prior). Across rounds 13 and 14, at ages 18 years and over, weighted prevalence of swab-positivity was 0.55% (0.50%, 0.61%) for those who received their second dose 3-6 months before their swab compared to 0.35% (0.31%, 0.40%) for those whose second dose was within 3 months of their swab, while weighted prevalence among unvaccinated individuals was1.76% (1.60%, 1.95%). In round 14, age group, region, key worker status, and household size jointly contributed to the risk of higher prevalence of swab-positivity.

**Discussion:** In September 2021 infections were increasing exponentially in the 5-to-17-year age group coinciding with the start of the autumn school term in England. Relatively few schoolchildren aged 5 to 17 years have been vaccinated in the UK though single doses are now being offered to those aged 12 years and over. In adults, the higher prevalence of swab-positivity following two doses of vaccine from 3 to 6 months compared to within 3 months of second dose supports the use of a booster vaccine. It is important that the vaccination programme maintains high coverage and reaches children and unvaccinated or partially vaccinated adults to reduce transmission and associated disruptions to work and education.

## Introduction

The United Kingdom (UK) has experienced one of the highest SARS-CoV-2 infection and COVID-19 fatality rates in Europe since the start of the pandemic [1]. However, the UK was one of the first to implement a national vaccination programme, starting in December 2020, with rollout to the population targeted at those most at risk including those at older ages, health care workers and people with specified health conditions.

The rapid spread of the Delta variant in England from May 2021 coincided with a third wave of infections [2] and prevalence of infections remained high into summer 2021 [3]. During August 2021 the incidence of reverse transcription-polymerase chain reaction (RT-PCR) confirmed cases of COVID-19 presenting to the national testing programme in England (Pillar 2) increased gradually by over 10% overall [3].

In September 2021 the rollout of the national vaccination programme against COVID-19 in England was extended to offer a single dose to older school children (aged 12 years and over) and booster (third) doses (at least six months following the second dose) to health and social care workers, all those over 50 years of age and younger people at risk. However, by mid-September 2021, the number of people receiving first vaccination doses dropped to its lowest level (just under 21,000 doses per day on average in the UK) [4] since at least mid-January 2021 (the earliest data available publicly). Nonetheless, by 12 October 2021, 85.6% of those 12 years of age and older in the UK had received their first dose and 78.6% had received their second [4].

In early September 2021, schoolchildren in England returned to school with the Department for Education no longer recommending that it was necessary to keep children in consistent groups (bubbles). Furthermore, schools were no longer expected to undertake contact tracing, with close contacts in schools now identified by the national contact tracing programme (Test and Trace) [5]. During this first month of return to school, the incidence of confirmed cases in England recorded through Pillar 2 dropped to a low in mid-September 2021 and then rose again, but overall was relatively stable. This is in stark contrast to the experience in Scotland where the incidence of confirmed cases increased by more than 450% during August, at a time when schoolchildren in Scotland returned to school [6]. Understanding such national-level (and more local) trends is highly dependent on the context, including return to school, changes in social mixing patterns, home versus office working and levels of vaccine-induced and naturally acquired immunity.

Here we describe the underlying dynamics driving patterns in SARS-CoV-2 during September 2021 in England by analysing RT-PCR swab-positivity data from the most recent round of the REal-time Assessment of Community Transmission-1 (REACT-1) study [7,8], round 14. This obtained throat and nose swabs from a random sample of the population of England at ages 5 years and over from 9 to 27 September 2021. We also combine round 14 data with data from round 13, which obtained swabs from 24 June to 12 July 2021.

## Results

### Descriptive statistics

A total of 822,176 participants were invited to participate in round 14 of the REACT-1 study of whom 147,393 (17.9%) registered and 100,527 (12.2%) provided a wet swab (using saline) with a valid result from RT-PCR. Samples were transported either by courier (N=46,705) or sent through the priority COVID-19 postal service (N=53,822) (Supplementary Table S1). Prevalence was slightly higher (Table S1) and cycle threshold (Ct) values in positives slightly lower (Table S1, Figure S1) in the samples sent by courier compared to samples shipped by post. Information on vaccination status and timing was available from the National Health Service (NHS) COVID-19 vaccine database for 87,966 (87.5%) participants who consented to data linkage.

### Prevalence and Epidemic Growth Estimates

Of the 100,527 valid swabs, a total of 764 were positive, giving a weighted prevalence of 0.83% (0.76%, 0.89%) (Table 1). This is higher than in round 13 when weighted prevalence was 0.63% (0.57%, 0.69%), despite the potential loss in diagnostic sensitivity due to the change in sample handling procedures in round 14.

**Table 1.** Unweighted and weighted prevalence of swab-positivity from REACT-1 across rounds 1 to 14

| Round | Tested swabs | Positive swabs | Unweighted prevalence (95% CI) | Weighted prevalence (95% CI) | First sample | Last sample |
| --- | --- | --- | --- | --- | --- | --- |
| 1 | 120,620 | 159 | 0.13% (0.11%, 0.15%) | 0.16% (0.13%, 0.19%) | 01/05/20 | 01/06/20 |
| 2 | 159,199 | 123 | 0.077% (0.065%, 0.092%) | 0.088% (0.068%, 0.11%) | 19/06/20 | 07/07/20 |
| 3 | 162,821 | 54 | 0.033% (0.025%, 0.043%) | 0.040% (0.027%, 0.053%) | 24/07/20 | 11/08/20 |
| 4 | 154,325 | 137 | 0.089% (0.075%, 0.11%) | 0.13% (0.096%, 0.15%) | 20/08/20 | 08/09/20 |
| 5 | 174,949 | 824 | 0.47% (0.44%, 0.50%) | 0.60% (0.55%, 0.71%) | 18/09/20 | 05/10/20 |
| 6 | 160,175 | 1,732 | 1.08% (1.03%, 1.13%) | 1.30% (1.21%, 1.39%) | 16/10/20 | 02/11/20 |
| 7 | 168,181 | 1,299 | 0.77% (0.73%, 0.82%) | 0.94% (0.87%, 1.01%) | 13/11/20 | 03/12/20 |
| 8 | 167,642 | 2,282 | 1.36% (1.31%, 1.42%) | 1.57% (1.49%, 1.66%) | 06/01/21 | 22/01/21 |
| 9 | 165,456 | 689 | 0.42% (0.39%, 0.45%) | 0.49% (0.44%, 0.55%) | 04/02/21 | 23/02/21 |
| 10 | 140,844 | 227 | 0.16% (0.14%, 0.18%) | 0.20% (0.17%, 0.23%) | 11/03/21 | 30/03/21 |
| 11 | 127,408 | 115 | 0.09% (0.07%, 0.11%) | 0.10% (0.08%, 0.13%) | 15/04/21 | 03/05/21 |
| 12* | 108,911 | 135 | 0.12% (0.10%, 0.15%) | 0.15% (0.12%, 0.18%) | 20/05/21 | 07/06/21 |
| 13 | 98,233 | 527 | 0.54% (0.49%, 0.58%) | 0.63% (0.57%, 0.69%) | 24/06/21 | 12/07/21 |
| 14** | 100,527 | 764 | 0.76% (0.71%, 0.82%) | 0.83% (0.76%, 0.89%) | 09/09/21 | 27/09/21*** |

\* Sampling strategy changed for round 12 and subsequent rounds. Therefore unweighted prevalence is not directly comparable with previous rounds
\*\* Sample handling changed in round 14. Therefore prevalence is not directly comparable with previous rounds
\*\*\* 509 (including 9 positive) additional samples (<0.5%) were received 28-30 September 2021 and are included in round 14

The P-spline fit to data from all REACT-1 rounds was indicative of a stable or increasing trend in the prevalence of swab-positivity during round 14 (Figure 1). A log-linear model estimated a reproduction number R of 1.03 (0.94, 1.14) across all age groups combined (Table 2). There were limited differences in the R estimates for samples shipped by post at 1.06 (0.93, 1.20) or by courier at 0.98 (0.84, 1.13) (Table 2).

**Table 2.**
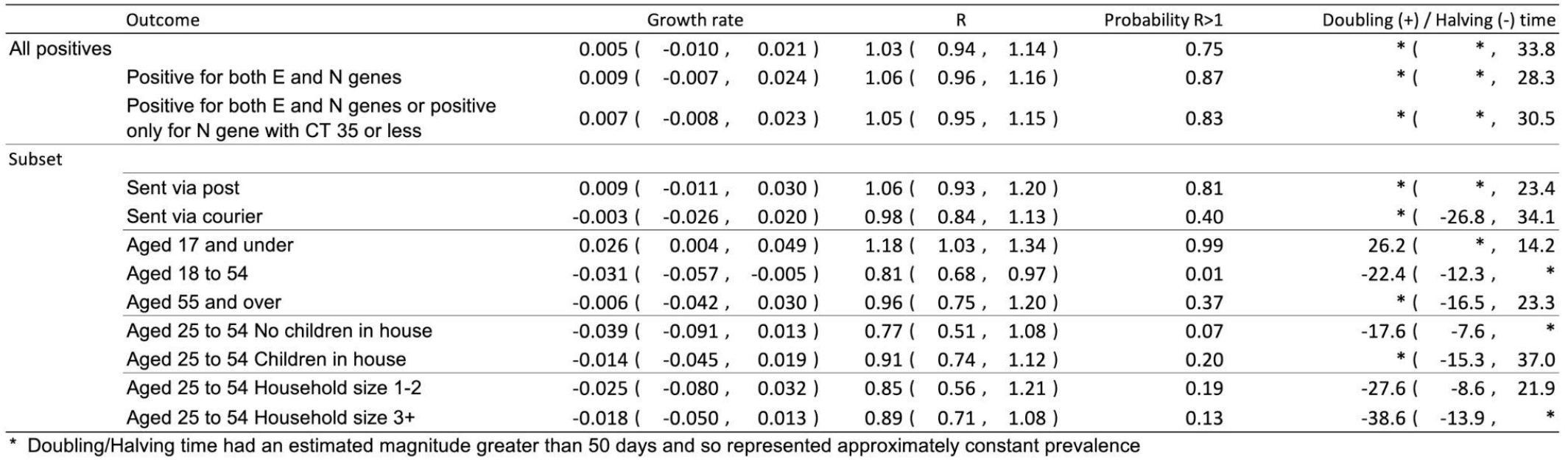
Table of growth rates, reproduction numbers and doubling/halving times from exponential model fits.

**Figure 1.**
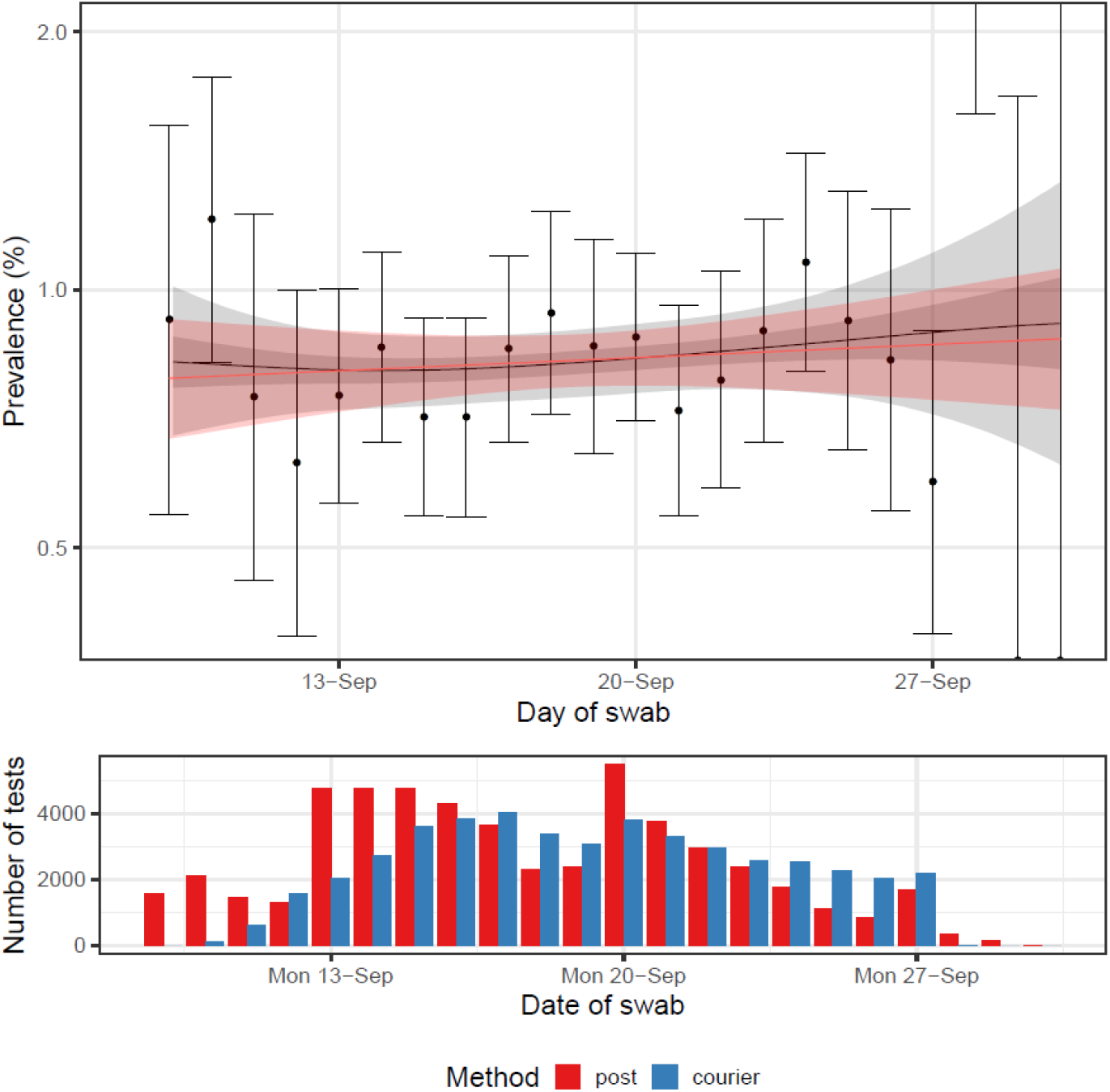
(Top) Comparison of an exponential model fit to round 14 (red) and a P-spline model fit to all rounds of REACT-1 (black, shown here only for round 14). Shaded red region shows the 95% posterior credible interval for the exponential model, and the shaded grey region shows 50% (dark grey) and 95% (light grey) posterior credible interval for the P-spline model. Results are presented for each day (X axis) of sampling for round 14 and the prevalence of swab-positivity is shown (Y axis) on a log scale. Weighted observations (black dots) and 95% confidence intervals (vertical lines) are also shown. (Bottom) Frequency of tests collected via courier (blue) and via post (red) by date of swab.

Weighted prevalence in round 14 varied by age group and ranged from 0.29% (0.20%, 0.42%) in adults aged 75 and over to 2.55% (2.11%, 3.08%) in teenagers aged 13 to 17. A high weighted prevalence at 2.32% (1.96%, 2.73%) was also observed in children aged 5 to 12 years (Figure 2, Table 3a).

**Table 3a.** Weighted prevalence of swab-positivity by sex, age, region, employment type, and ethnic group for round 14.

| Variable | Category | Round 14 |  |  |  |
| --- | --- | --- | --- | --- | --- |
|  |  | Positive | Total | Unweighted Prevalence | Weighted Prevalence* |
| Gender | Male | 336 | 44,549 | 0.75% ( 0.68% , 0.84% ) | 0.82% ( 0.73% , 0.92% ) |
|  | Female | 428 | 55,976 | 0.76% ( 0.69% , 0.84% ) | 0.83% ( 0.75% , 0.93% ) |
|  | Unknown | 0 | 2 | 0.00% ( 0.00% , 84.19% ) | 0.00% ( 0.00% , 0.00% ) |
| Age | 05-12 | 155 | 6,458 | 2.40% ( 2.04% , 2.80% ) | 2.32% ( 1.96% , 2.73% ) |
|  | 13-17 | 118 | 4,927 | 2.40% ( 1.99% , 2.86% ) | 2.55% ( 2.11% , 3.08% ) |
|  | 18-24 | 9 | 2,452 | 0.37% ( 0.17% , 0.70% ) | 0.46% ( 0.23% , 0.90% ) |
|  | 25-34 | 28 | 7,374 | 0.38% ( 0.25% , 0.55% ) | 0.36% ( 0.24% , 0.53% ) |
|  | 35-44 | 100 | 12,118 | 0.83% ( 0.67% , 1.00% ) | 0.79% ( 0.64% , 0.97% ) |
|  | 45-54 | 130 | 16,855 | 0.77% ( 0.64% , 0.92% ) | 0.78% ( 0.65% , 0.93% ) |
|  | 55-64 | 113 | 20,856 | 0.54% ( 0.45% , 0.65% ) | 0.55% ( 0.45% , 0.67% ) |
|  | 65-74 | 80 | 19,313 | 0.41% ( 0.33% , 0.52% ) | 0.42% ( 0.34% , 0.53% ) |
|  | 75+ | 31 | 10,174 | 0.30% ( 0.21% , 0.43% ) | 0.29% ( 0.20% , 0.42% ) |
| Region | South East | 92 | 17,388 | 0.53% ( 0.43% , 0.65% ) | 0.57% ( 0.45% , 0.72% ) |
|  | North East | 39 | 4,551 | 0.86% ( 0.61% , 1.17% ) | 0.84% ( 0.60% , 1.18% ) |
|  | North West | 121 | 12,117 | 1.00% ( 0.83% , 1.19% ) | 0.99% ( 0.81% , 1.21% ) |
|  | Yorkshire and The Humber | 105 | 9,887 | 1.06% ( 0.87% , 1.28% ) | 1.25% ( 1.00% , 1.57% ) |
|  | East Midlands | 87 | 8,830 | 0.99% ( 0.79% , 1.21% ) | 1.15% ( 0.92% , 1.44% ) |
|  | West Midlands | 88 | 10,249 | 0.86% ( 0.69% , 1.06% ) | 1.01% ( 0.80% , 1.27% ) |
|  | East of England | 87 | 11,756 | 0.74% ( 0.59% , 0.91% ) | 0.73% ( 0.59% , 0.92% ) |
|  | London | 91 | 14,885 | 0.61% ( 0.49% , 0.75% ) | 0.62% ( 0.50% , 0.79% ) |
|  | South West | 54 | 10,864 | 0.50% ( 0.37% , 0.65% ) | 0.59% ( 0.43% , 0.80% ) |
| Employment type | Health care or care home worker | 59 | 7,963 | 0.74% ( 0.56% , 0.95% ) | 0.80% ( 0.60% , 1.06% ) |
|  | Other essential/key worker | 159 | 14,627 | 1.09% ( 0.93% , 1.27% ) | 1.07% ( 0.90% , 1.28% ) |
|  | Other worker | 263 | 38,496 | 0.68% ( 0.60% , 0.77% ) | 0.71% ( 0.62% , 0.82% ) |
|  | Not full-time, part-time, or self-employed | 244 | 37,312 | 0.65% ( 0.57% , 0.74% ) | 0.77% ( 0.67% , 0.89% ) |
|  | Unknown | 39 | 2,129 | 1.83% ( 1.31% , 2.50% ) | 1.88% ( 1.34% , 2.64% ) |
| Ethnic group | White | 635 | 87,942 | 0.72% ( 0.67% , 0.78% ) | 0.78% ( 0.72% , 0.85% ) |
|  | Asian | 58 | 5,550 | 1.05% ( 0.79% , 1.35% ) | 1.04% ( 0.77% , 1.41% ) |
|  | Black | 22 | 1,947 | 1.13% ( 0.71% , 1.71% ) | 1.41% ( 0.91% , 2.19% ) |
|  | Mixed | 18 | 1,754 | 1.03% ( 0.61% , 1.62% ) | 1.01% ( 0.62% , 1.63% ) |
|  | Other | 12 | 1,015 | 1.18% ( 0.61% , 2.06% ) | 1.01% ( 0.56% , 1.82% ) |
|  | Unknown | 19 | 2,319 | 0.82% ( 0.49% , 1.28% ) | 1.09% ( 0.67% , 1.75% ) |

\* For categories other than age and region, we present weighted prevalence if the number of positives in a category is 10 or more.

**Table 3b.**
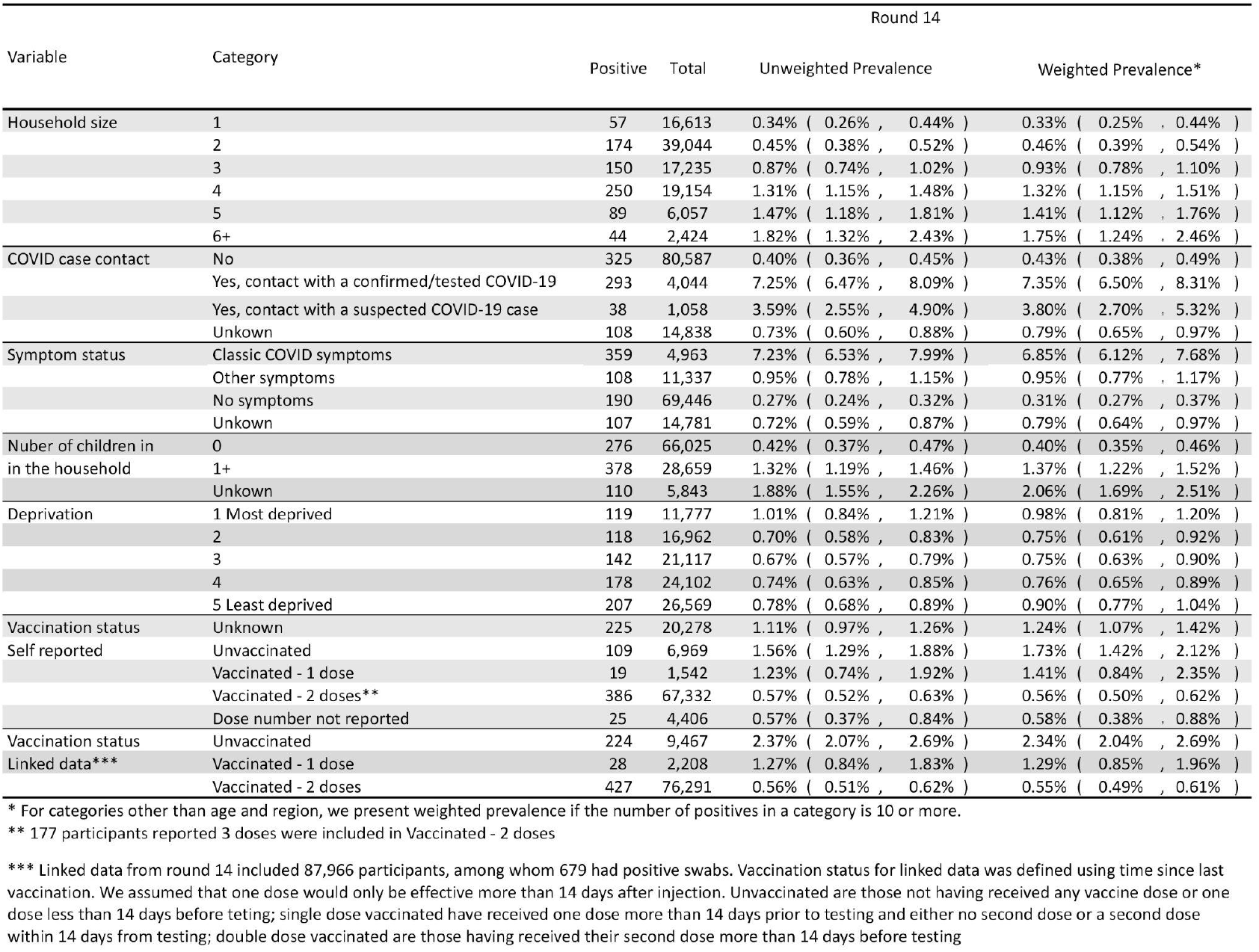
Weighted prevalence of swab-positivity by household size, COVID-19 case contact status, symptom status, number of children in the household, neighbourhood deprivation and vaccination status for round 14.

**Figure 2.**
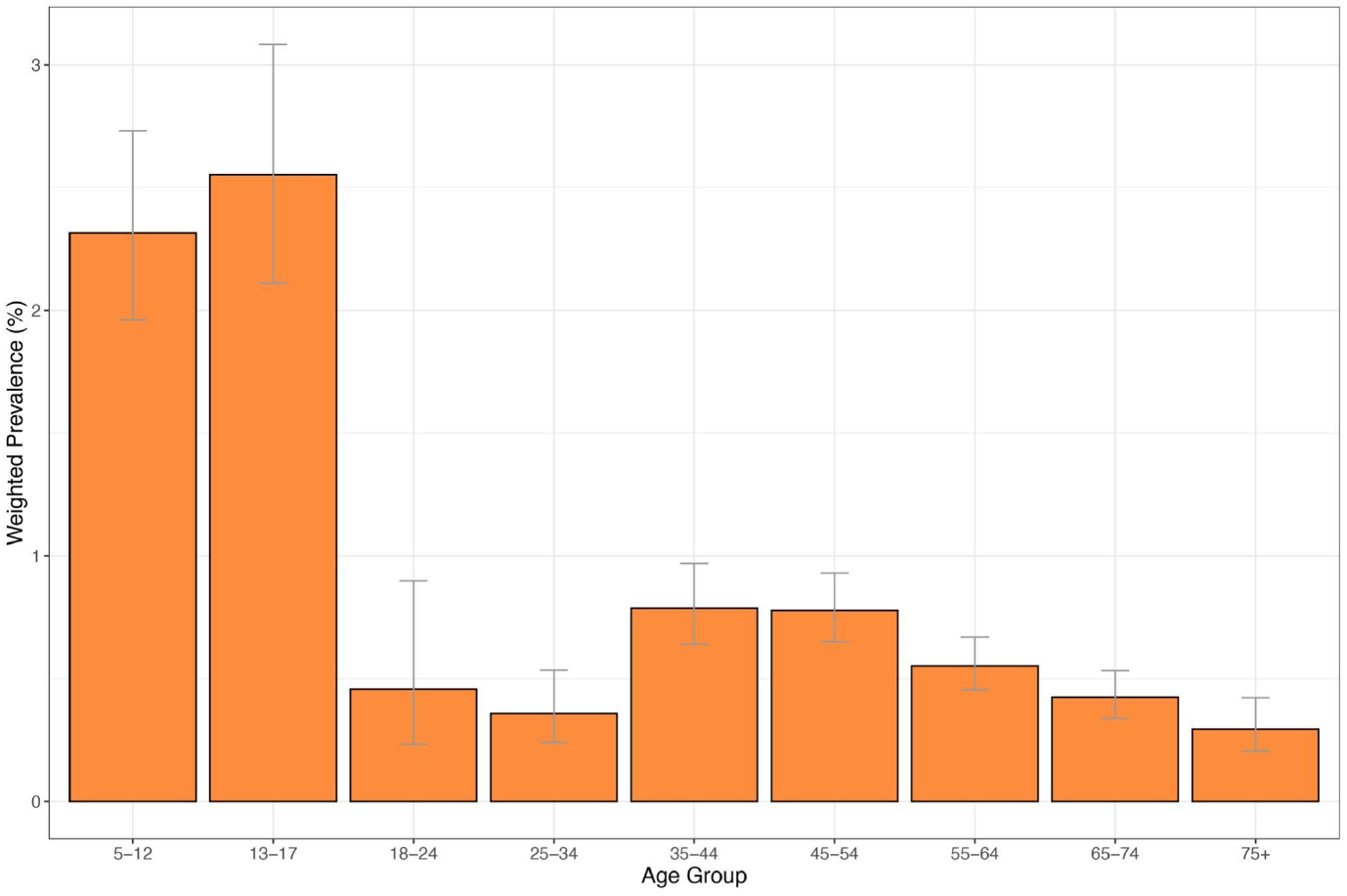
Weighted prevalence of swab-positivity by age group in round 14. Bars show the prevalence point estimates, and the vertical lines represent the 95% confidence intervals.

The fitted P-splines indicated increasing prevalence at ages 5 to 17 years and decreasing prevalence at ages 18 to 54 years (Figure 3). Log-linear models estimated a >0.99 posterior probability that the growth rate differed between these two groups. Accordingly, we estimated that the epidemic was growing in round 14 among those aged 17 years and under with R of 1.18 (1.03, 1.34) and 0.99 posterior probability that R>1, and decreasing with R of 0.81 (0.68, 0.97) and 0.01 posterior probability that R>1, in those aged 18-54 years (Table 2).

**Figure 3.**
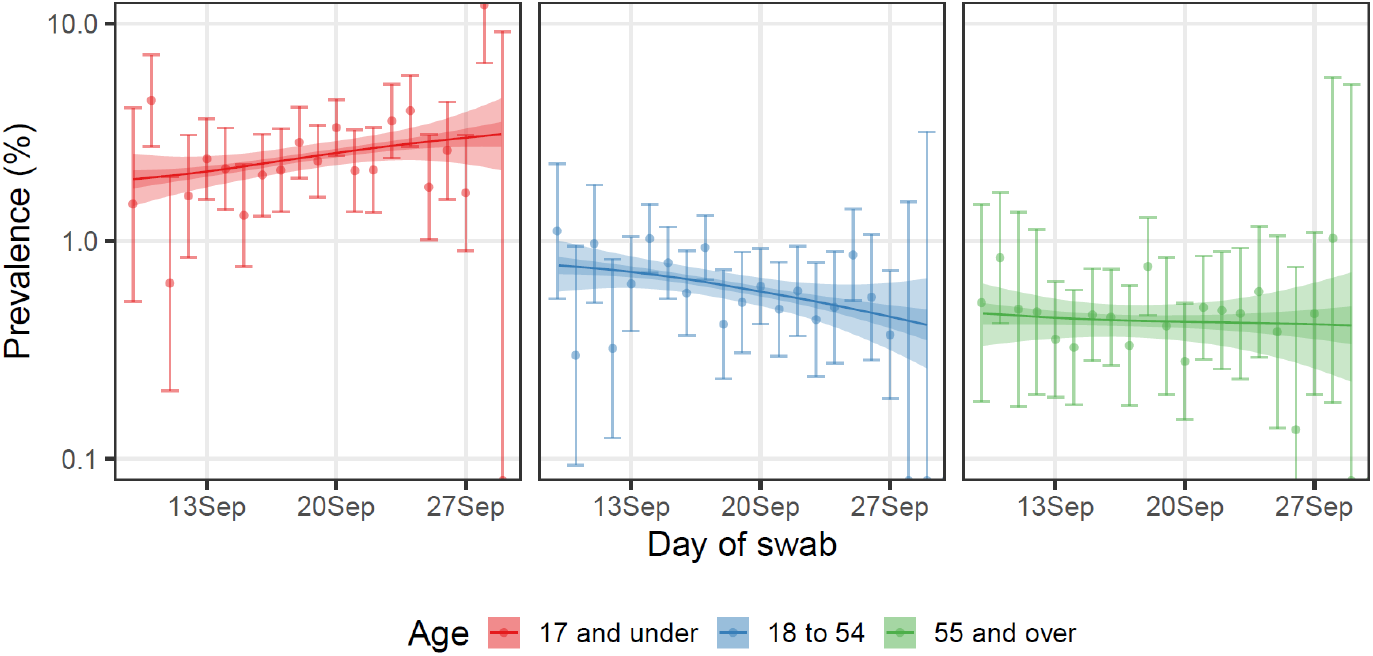
Comparison of P-spline models fit to all rounds of REACT-1 for those aged 17 and under (red), those aged 18 to 54 inclusive (blue) and those aged 55 and over (green). Shown here for only the period of round 14. Shaded regions show 50% (dark shade) and 95% (light shade) posterior credible interval for the P-spline models. Results are presented for each day (X axis) of sampling for round 14 and the prevalence of swab-positivity is shown (Y axis) on a log scale. Weighted observations (dots) and 95% confidence intervals (vertical lines) are also shown.

Weighted prevalence was also found to vary by region (Figure 4, Table 3a) ranging from 0.57% (0.45%, 0.72%) in the South East to 1.25% (1.00%, 1.57%) in Yorkshire and The Humber. Within round 14, there was evidence of epidemic growth (with posterior probability that R>1 greater than or equal to 0.99) in London with R of 1.59 (1.23, 1.99) and in East Midlands with R of 1.36 (1.05, 1.73) (Table 4).

**Table 4.**
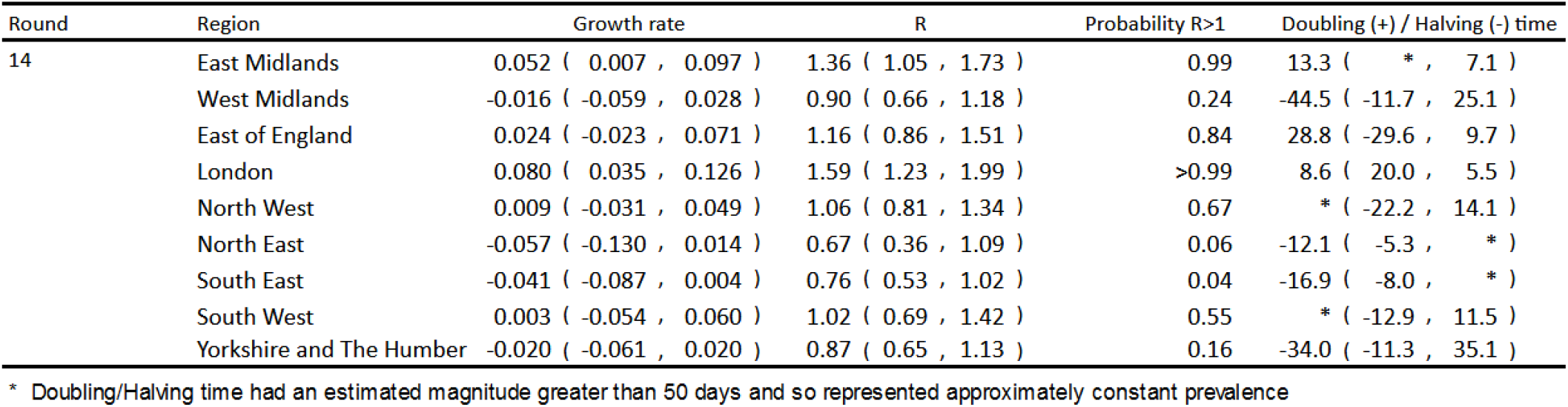
Table of estimated growth rates, reproduction numbers and doubling/halving times from exponential model fits by region for round 14.

| Round | Region | Growth rate | R | Probability R>1 | Doubling (+) / Halving (-) time |
| --- | --- | --- | --- | --- | --- |
| 14 | East Midlands | 0.052 ( 0.007 , 0.097 ) | 1.36 ( 1.05 , 1.73 ) | 0.99 | 13.3 ( * , 7.1 ) |
|  | West Midlands | -0.016 ( -0.059 , 0.028 ) | 0.90 ( 0.66 , 1.18 ) | 0.24 | -44.5 ( -11.7 , 25.1 ) |
|  | East of England | 0.024 ( -0.023 , 0.071 ) | 1.16 ( 0.86 , 1.51 ) | 0.84 | 28.8 ( -29.6 , 9.7 ) |
|  | London | 0.080 ( 0.035 , 0.126 ) | 1.59 ( 1.23 , 1.99 ) | >0.99 | 8.6 ( 20.0 , 5.5 ) |
|  | North West | 0.009 ( -0.031 , 0.049 ) | 1.06 ( 0.81 , 1.34 ) | 0.67 | * ( -22.2 , 14.1 ) |
|  | North East | -0.057 ( -0.130 , 0.014 ) | 0.67 ( 0.36 , 1.09 ) | 0.06 | -12.1 ( -5.3 , * ) |
|  | South East | -0.041 ( -0.087 , 0.004 ) | 0.76 ( 0.53 , 1.02 ) | 0.04 | -16.9 ( -8.0 , * ) |
|  | South West | 0.003 ( -0.054 , 0.060 ) | 1.02 ( 0.69 , 1.42 ) | 0.55 | * ( -12.9 , 11.5 ) |
|  | Yorkshire and The Humber | -0.020 ( -0.061 , 0.020 ) | 0.87 ( 0.65 , 1.13 ) | 0.16 | -34.0 ( -11.3 , 35.1 ) |

\* Doubling/Halving time had an estimated magnitude greater than 50 days and so represented approximately constant prevalence

**Figure 4.**
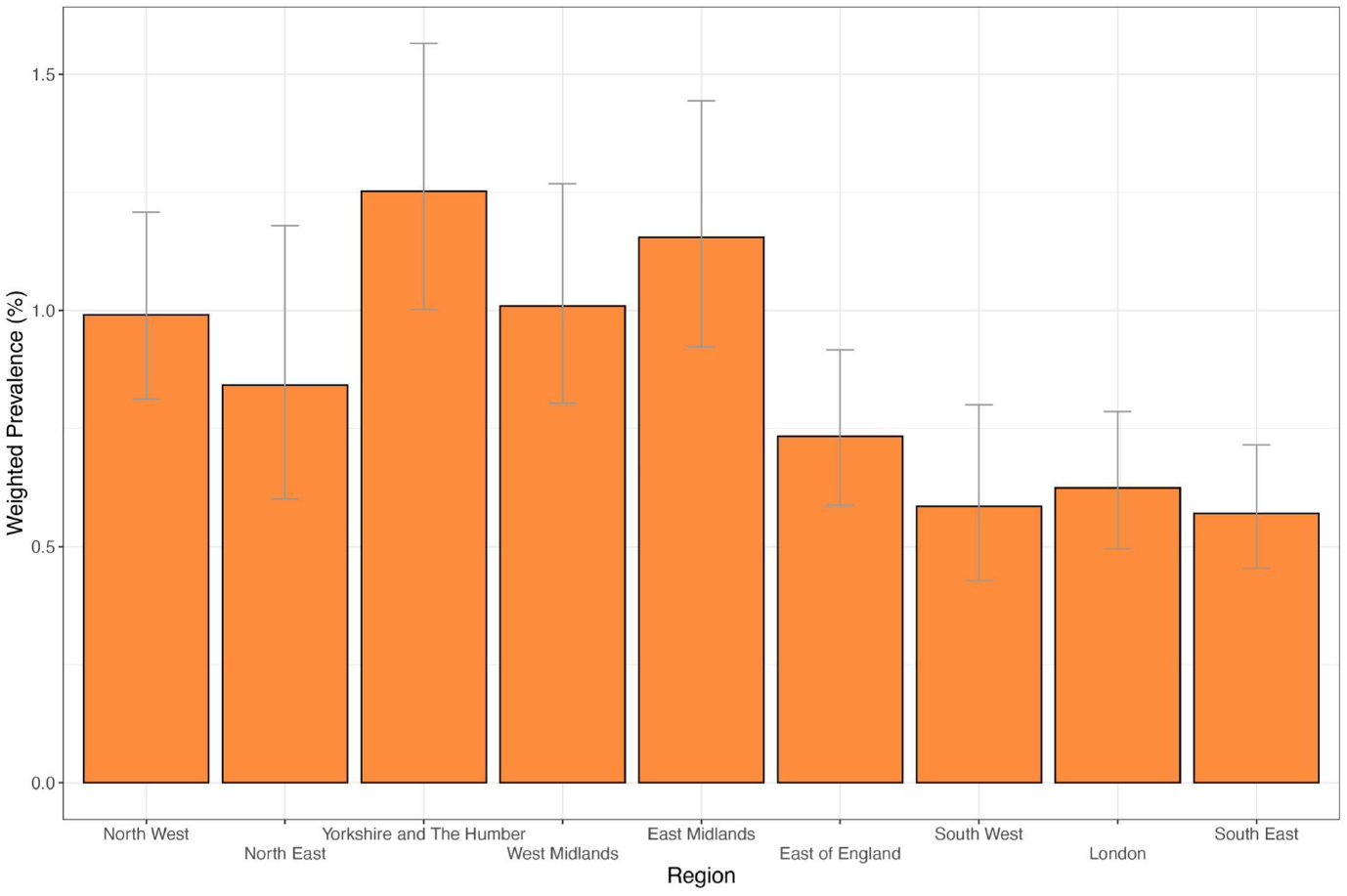
Weighted prevalence of swab-positivity by region in round 14. Bars represent prevalence point estimates, and the vertical lines the corresponding 95% confidence intervals.

We found higher weighted prevalence of swab-positivity among participants of Black ethnicity at 1.41% (0.91%, 2.19%), compared to that of white participants at 0.78% (0.72%, 0.85%) (Table 3a). Participants living in larger compared to smaller households had higher weighted prevalence ranging from 0.33% (0.25%, 0.44%) for single-person households to 1.75% (1.24%, 2.46%) for households with 6 or more persons (Table 3b). Prevalence was also higher in households with one or more children at 1.37% (1.22%, 1.52%) compared to 0.40% (0.35%, 0.46%) for households without children; and among those reporting to have been in contact with a confirmed COVID-19 case at 7.35% (6.50%, 8.31%) compared with 0.43% (0.38%, 0.49%) among those without such contact.

Using self-reported vaccination status we found higher prevalence in unvaccinated participants at 1.73% (1.42%, 2.12%) at all ages compared to those reporting having received two vaccine doses at 0.56% (0.50%, 0.62%) (Table 3b). To account for time needed for the vaccine to grant immunity, we recoded the vaccination status using time since last vaccination assuming that a dose (first and second) would only be effective more than 14 days after injection. Due to the large number of missing dates in self-reported data, we used linked data for which information on dates was almost complete. Using this definition of vaccination status on the linked data, we found a similar gradient in weighted prevalence ranging from 0.55% (0.49%, 0.61%) in those having received two doses to 2.34% (2.04%, 2.69%) in unvaccinated people.

In multiple logistic regression analysis (Table 5) key workers other than healthcare workers and care home workers had increased risk of swab-positivity, with odds ratio (OR) of 1.35 (1.10, 1.66) compared to other workers. Swab-positivity was also higher with increasing household size, with mutually adjusted ORs of 1.77 (1.44, 2.17) and 2.37 (1.62, 3.47) for households of 3-5 and 6 or more persons, respectively, compared to households with 1 or 2 persons.

**Table 5.**
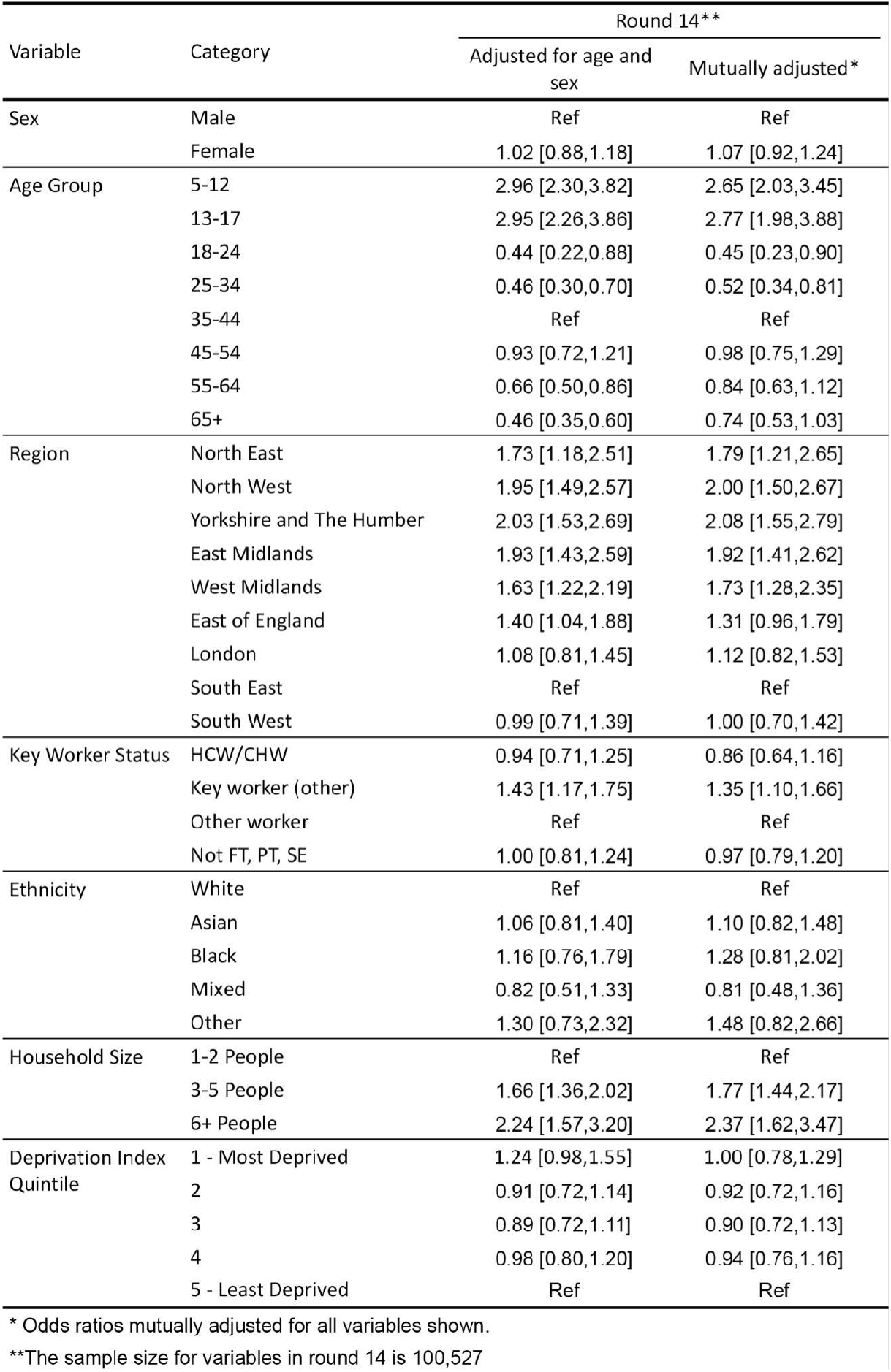
Multiple logistic regression for round 14. Results are presented as odds ratios (ORs) and 95% confidence intervals adjusted for age and sex and additionally for all other variables (mutually adjusted OR).

| Variable | Category | Round 14** |  |
| --- | --- | --- | --- |
|  |  | Adjusted for age and sex | Mutually adjusted* |
| Sex | Male | Ref | Ref |
|  | Female | 1.02 [0.88,1.18] | 1.07 [0.92,1.24] |
| Age Group | 5-12 | 2.96 [2.30,3.82] | 2.65 [2.03,3.45] |
|  | 13-17 | 2.95 [2.26,3.86] | 2.77 [1.98,3.88] |
|  | 18-24 | 0.44 [0.22,0.88] | 0.45 [0.23,0.90] |
|  | 25-34 | 0.46 [0.30,0.70] | 0.52 [0.34,0.81] |
|  | 35-44 | Ref | Ref |
|  | 45-54 | 0.93 [0.72,1.21] | 0.98 [0.75,1.29] |
|  | 55-64 | 0.66 [0.50,0.86] | 0.84 [0.63,1.12] |
|  | 65+ | 0.46 [0.35,0.60] | 0.74 [0.53,1.03] |
| Region | North East | 1.73 [1.18,2.51] | 1.79 [1.21,2.65] |
|  | North West | 1.95 [1.49,2.57] | 2.00 [1.50,2.67] |
|  | Yorkshire and The Humber | 2.03 [1.53,2.69] | 2.08 [1.55,2.79] |
|  | East Midlands | 1.93 [1.43,2.59] | 1.92 [1.41,2.62] |
|  | West Midlands | 1.63 [1.22,2.19] | 1.73 [1.28,2.35] |
|  | East of England | 1.40 [1.04,1.88] | 1.31 [0.96,1.79] |
|  | London | 1.08 [0.81,1.45] | 1.12 [0.82,1.53] |
|  | South East | Ref | Ref |
|  | South West | 0.99 [0.71,1.39] | 1.00 [0.70,1.42] |
| Key Worker Status | HCW/CHW | 0.94 [0.71,1.25] | 0.86 [0.64,1.16] |
|  | Key worker (other) | 1.43 [1.17,1.75] | 1.35 [1.10,1.66] |
|  | Other worker | Ref | Ref |
|  | Not FT, PT, SE | 1.00 [0.81,1.24] | 0.97 [0.79,1.20] |
| Ethnicity | White | Ref | Ref |
|  | Asian | 1.06 [0.81,1.40] | 1.10 [0.82,1.48] |
|  | Black | 1.16 [0.76,1.79] | 1.28 [0.81,2.02] |
|  | Mixed | 0.82 [0.51,1.33] | 0.81 [0.48,1.36] |
|  | Other | 1.30 [0.73,2.32] | 1.48 [0.82,2.66] |
| Household Size | 1-2 People | Ref | Ref |
|  | 3-5 People | 1.66 [1.36,2.02] | 1.77 [1.44,2.17] |
|  | 6+ People | 2.24 [1.57,3.20] | 2.37 [1.62,3.47] |
| Deprivation Index Quintile | 1 - Most Deprived | 1.24 [0.98,1.55] | 1.00 [0.78,1.29] |
|  | 2 | 0.91 [0.72,1.14] | 0.92 [0.72,1.16] |
|  | 3 | 0.89 [0.72,1.11] | 0.90 [0.72,1.13] |
|  | 4 | 0.98 [0.80,1.20] | 0.94 [0.76,1.16] |
|  | 5 - Least Deprived | Ref | Ref |

\* Odds ratios mutually adjusted for all variables shown.
\*\*The sample size for variables in round 14 is 100,527

### Vaccination status and Vaccine Effectiveness

Pooling the linked data from rounds 13 and 14 for ages 18 years and over (in total N=172,862), we investigated weighted prevalence of swab-positivity by vaccination status (Figure 5). Across rounds and age group (for those over 18 years), weighted prevalence was higher at 0.55% (0.50%, 0.61%) for those who received their second dose 3-6 months before their swab compared to 0.35% (0.31%, 0.40%) for those whose second dose was within 3 months of the swab (Figure 5A). The weighted prevalence for those whose second dose was administered more than 6 months prior was similar to those vaccinated within 3-6 months of their swab at 0.52% (0.33%, 0.78%), but with a wider confidence interval. Nevertheless, irrespective of the number and timing of the vaccine doses received, weighted prevalence in vaccinated people at ages 18 years and over was lower than that in unvaccinated people, which was 1.76% (1.60%, 1.95%). The age distribution across vaccine categories (Figure 5B) was heterogeneous, with a higher proportion of younger participants being unvaccinated or having received only a single vaccine dose. However, in the double-vaccinated participants there was an apparent trend of increasing weighted prevalence at each age for those having received their second dose 3-6 months before their swab compared to those vaccinated less than 3 months earlier (Figure 5C).

**Figure 5.**
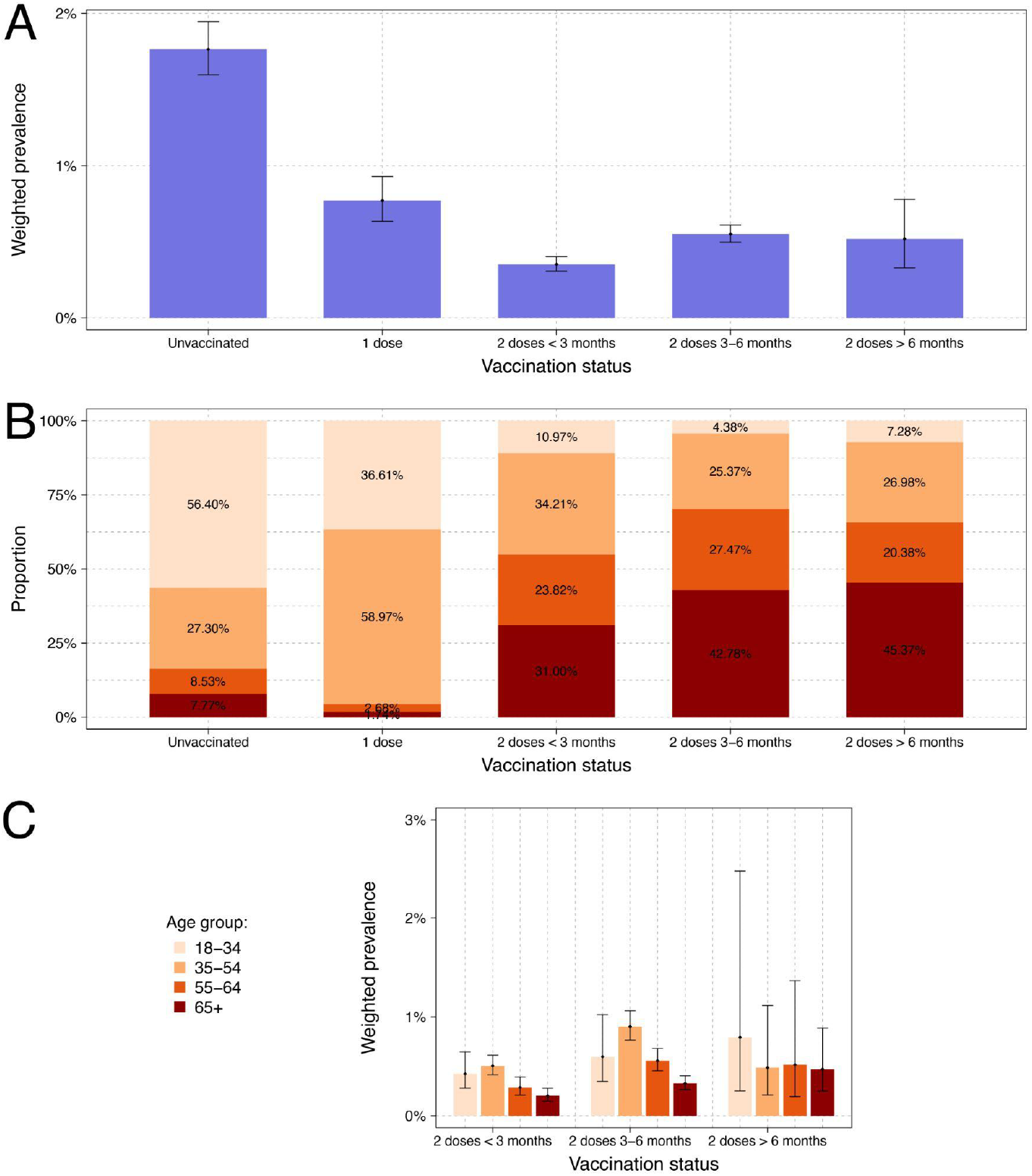
Weighted prevalence of swab-positivity for all REACT-1 participants aged 18 years and over with linked data in round 13 and round 14 combined by vaccination status (A). Age distribution within each vaccination status (B). Weighted prevalence by age group in participants who received two vaccine doses (C).

Vaccine Effectiveness (VE) estimates were derived using the linked dataset for rounds 13 and 14 combined restricted to ages 18 to 64 years only, since few people were unvaccinated over the age of 65 years, and few were vaccinated below the age of 18 years. VE was estimated through a series of logistic models adjusted for round, and sequentially for age, sex, and additionally for index of multiple deprivation, region and ethnicity (Table 6) (all Delta variant infections, see below). In the full population at ages 18 to 64 years and for all vaccines combined, VE against infection ranged from 66.3% (55.3%, 74.7%) with adjustment for round to 62.8% (49.3%, 72.7%) in the fully adjusted model. Fully adjusted VE estimates for AstraZeneca and Pfizer-BioNTech vaccines were 44.8% (22.5%, 60.7%) and 71.3% (56.6%, 81.0%), respectively. VE estimates in models restricted to symptomatic cases were similar to those of models that also included asymptomatic cases.

**Table 6.** Estimates of vaccine effectiveness against swab-positivity indicative of breakthrough infections for round 13 and 14 of REACT-1 adjusted for round and further adjusted for age and sex, and IMD, region, and ethnicity: for linked vaccine status data, for participants aged 18 to 64 years and test positive reporting at least one symptom in month prior to testing.

|  | Vaccination Status | Adjustment | Test Negatives |  |  | Test Positives |  |  | Vaccine Effectiveness (95% Confidence Interval) | Interaction pvalue* |
| --- | --- | --- | --- | --- | --- | --- | --- | --- | --- | --- |
|  |  |  | Round 13 | Round 14 | Total | Round13 | Round 14 | Total |  |  |
| Full population | unvaccinated |  | 2907 | 1026 | 3933 | 44 | 13 | 57 |  |  |
|  | All vaccines | Round | 34853 | 49688 | 84541 | 144 | 328 | 472 | 66.3% ( 55.3% , 74.7% ) | 0.05 |
|  |  | Round, Age, Sex |  |  |  |  |  |  | 61.4% ( 47.5% , 71.6% ) | 0.11 |
|  |  | Round, Age, Sex, IMD, Region, Ethnicity |  |  |  |  |  |  | 62.8% ( 49.3% , 72.7% ) | 0.90 |
|  | AZ | Round | 24934 | 31290 | 56224 | 109 | 248 | 357 | 62.3% ( 49.7% , 71.8% ) | 0.02 |
|  |  | Round, Age, Sex |  |  |  |  |  |  | 41.8% ( 18.5% , 58.4% ) | 0.13 |
|  |  | Round, Age, Sex, IMD, Region, Ethnicity |  |  |  |  |  |  | 44.8% ( 22.5% , 60.7% ) | 0.11 |
|  | Pfizer | Round | 6500 | 12888 | 19388 | 26 | 56 | 82 | 70.7% ( 57.5% , 79.7% ) | 0.51 |
|  |  | Round, Age, Sex |  |  |  |  |  |  | 70.0% ( 55.0% , 80.0% ) | 0.56 |
|  |  | Round, Age, Sex, IMD, Region, Ethnicity |  |  |  |  |  |  | 71.3% ( 56.6% , 81.0% ) | 0.49 |
|  | Moderna | Round | 24 | 1197 | 1221 | 0 | 4 | 4 | 74.4% ( 21.9% , 91.6% ) | 0.98 |
|  |  | Round, Age, Sex |  |  |  |  |  |  | 76.4% ( 27.6% , 92.3% ) | 0.98 |
|  |  | Round, Age, Sex, IMD, Region, Ethnicity |  |  |  |  |  |  | 75.1% ( 22.7% , 92.0% ) | 0.98 |
|  | Unknown | Round | 3395 | 4313 | 7708 | 9 | 20 | 29 | 74.9% ( 59.8% , 84.3% ) | 0.15 |
|  |  | Round, Age, Sex |  |  |  |  |  |  | 68.1% ( 46.6% , 81.0% ) | 0.28 |
|  |  | Round, Age, Sex, IMD, Region, Ethnicity |  |  |  |  |  |  | 67.1% ( 44.1% , 80.6% ) | 0.16 |
| Symptomatic positives only** | unvaccinated |  | 2907 | 1026 | 3933 | 44 | 13 | 32 |  |  |
|  | All vaccines | Round | 34853 | 49688 | 84541 | 64 | 224 | 288 | 67.8% ( 53.0% , 77.9% ) | 0.03 |
|  |  | Round, Age, Sex |  |  |  |  |  |  | 64.4% ( 46.8% , 76.2% ) | 0.04 |
|  |  | Round, Age, Sex, IMD, Region, Ethnicity |  |  |  |  |  |  | 66.4% ( 49.6% , 77.6% ) | 0.04 |
|  | AZ | Round | 24934 | 31290 | 56224 | 49 | 177 | 226 | 62.3% ( 44.9% , 74.3% ) | 0.01 |
|  |  | Round, Age, Sex |  |  |  |  |  |  | 42.4% ( 11.0% , 62.7% ) | 0.05 |
|  |  | Round, Age, Sex, IMD, Region, Ethnicity |  |  |  |  |  |  | 45.5% ( 15.5% , 64.9% ) | 0.06 |
|  | Pfizer | Round | 6500 | 12888 | 19388 | 9 | 33 | 42 | 76.5% ( 61.1% , 85.8% ) | 0.23 |
|  |  | Round, Age, Sex |  |  |  |  |  |  | 74.2% ( 55.4% , 85.1% ) | 0.34 |
|  |  | Round, Age, Sex, IMD, Region, Ethnicity |  |  |  |  |  |  | 76.6% ( 59.0% , 86.6% ) | 0.32 |
|  | Moderna | Round | 24 | 1197 | 1221 |  |  | 0 |  |  |
|  |  | Round, Age, Sex |  |  |  |  |  |  |  |  |
|  |  | Round, Age, Sex, IMD, Region, Ethnicity |  |  |  |  |  |  |  |  |
|  | Unknown | Round | 3395 | 4313 | 7708 | 6 | 14 | 20 | 70.2% ( 46.3% , 83.4% ) | 0.30 |
|  |  | Round, Age, Sex |  |  |  |  |  |  | 59.9% ( 23.2% , 79.1% ) | 0.53 |
|  |  | Round, Age, Sex, IMD, Region, Ethnicity |  |  |  |  |  |  | 61.1% ( 23.7% , 80.2% ) | 0.36 |

\* p-value for the multiplicative interaction term measuring possible modification of the effect of vaccination status by round
\*\* Symptomatic positives are defined as those with a positive swab having reported at least one of the 26 assayed symptoms in the month prior to testing

### Sequencing data

Lineage from the 475 (62.2%) sequenced among the 764 positive swabs all corresponded to Delta variant or sub-lineages of Delta (Table S2). AY.4 was the most detected sub-lineage representing 61.5% (57.0%, 65.7%) of the sequenced samples, and there were 7 samples with single spike mutations of interest (as defined by Public Health England): E484K, N501Y, F490S, R246G, V483F, P251L and Q613H.

## Discussion

In this fourteenth round of the REACT-1 study we found both high and increasing prevalence of SARS-CoV-2 swab-positivity among school-aged children in September 2021, reflecting increased social mixing of children as they attended school for the autumn term. At the same time we found decreasing prevalence among young to middle-aged adults (18 to 54 years). This may reflect the effect of previous natural infection, especially among younger adults where infection rates have been high [9], and the continued roll out of the vaccination campaign in England. Since April 2021 this was expanded to include adults under the age of 50 years [10], older teenagers (aged 16 and 17 years) from August 2021 and most recently (mid-September 2021) children aged 12 to 15 years.

Vaccination has proved highly effective against severe complications of COVID-19 including hospitalization and death, but there is less clarity concerning protection against infection. Nonetheless, estimates of VE against infection of up to 90% have been reported, although based on routine testing of symptomatic individuals [11]. We reported in round 13 of REACT-1 an estimate of VE against infection of 62% based on the linked data [12]. However, we were not powered to examine VE by vaccine type in round 13 alone. Here, we combined data from rounds 13 and 14 with an estimate of ca. 63% for VE overall, similar to our previous estimate. But we were also able to estimate VE for AstraZeneca and Pfizer-BioNTech vaccines separately. Though confidence intervals overlap, our results suggest higher effectiveness against infection for Pfizer-BioNTech than AstraZeneca, consistent with findings from Public Health England that reported higher VE for Pfizer-BioNTech based on routine testing of symptomatic cases (Pillar 2 of the national testing programme) [13]. However, it should be noted that VE is population- and time-specific so these estimates are context specific reflecting performance of the vaccines in England at this time.

As in our previous report (round 13 [12]), we found that all sequenced swabs were Delta variant and its sublineages, indicating almost complete replacement of Alpha and other variants by Delta in England. We detected one potential escape E484K mutation which translates into an estimated 984 such infections in England with a lower 95% confidence limit of 159. Overall there were 7 samples with single spike mutations of interest as defined by Public Health England (now part of the UK Health Security Agency).

Our study shows that prevalence of swab-positivity among unvaccinated individuals remains three to four-times higher than in double-vaccinated people, but also suggests that prevalence of swab-positivity indicative of breakthrough infections following two-dose vaccination may increase after 3 to 6 months. In addition, we find that people living in larger households, and people living in households with children, experience higher rates of swab-positivity than those in smaller households or without children in the household. Thus there is a delicate balance between degree of social mixing including across generations, levels of vaccination in the community and risk of infection.

Although following the vaccination campaign there has been a relative uncoupling between infections and hospitalizations and deaths in England [2], concerns remain about the potential for high infection rates and incomplete population immunity to result in increased risk of severe complications from COVID-19. England in common with the rest of the United Kingdom and several other countries (notably Israel [14]) has embarked on a campaign to roll out third (booster) doses (in England this involves adults aged 50 years and over, health and social care workers and younger people at risk) [15]. On-going monitoring of the epidemic in England and elsewhere will be important to gauge the extent to which booster doses in adults and the campaign to vaccinate older children curtail any future wave of the epidemic.

The finding of both the highest weighted prevalence levels and exponential growth in swab-positivity during round 14 in the 5-to-17-year age group raises concerns for clinically extremely vulnerable children as well as clinically extremely vulnerable close contacts including household members and school staff. There are also concerns about the effects on education of the large numbers of children who are required to be out of school when testing positive as a result of the high rates of infection. The finding that prevalence was more than three-times greater for individuals in households with one or more children than in those in households without children suggests that infections in children, unsurprisingly, spread into other age groups. Nonetheless, as noted, the declining prevalence of swab-positivity during round 14 in adults aged 18 to 54 years, suggests that these adults are benefitting from both high previous levels of natural infection (especially among younger adults) and the high uptake of vaccination (especially among older adults in this age range).

Our study has limitations. Since the REACT-1 study began in May 2020, we observed a gradual reduction in response rates from 30.5% in round 1 to 11.7% in round 13. However, in round 14, the response rate increased slightly to 12.2%. This increase, albeit small, is encouraging. It may be that further changes to the survey could further increase participation. The change to using wet swabs in saline solution and the collection of samples without the cold chain (by post or courier) may well have affected diagnostic sensitivity. We were reassured by the limited differences between the samples collected by post and courier. However, because the exact system used in previous rounds (dry swabs and a sustained cold chain) was not compared within a round, we could not estimate the impact of the new approaches compared to that used in previous rounds. A further limitation is that we do not have perfect data on the vaccination status of all participants. Although consent to data linkage was at a high level (87.5% in round 14) not all participants consent for linkage to their NHS records which include data from the COVID-19 immunization programme. For those who are not linked, data on the dates of vaccination and vaccine type are either missing or less reliable than in the linked data, such that we based our estimates of VE on the subset with linked records. This may introduce a bias to the extent that those who do and do not consent to data linkage may differ in important ways, such as social mixing patterns, that may affect risk of infection.

In addition, for our estimates of VE by vaccine type, even though we controlled for age and round in our sample, the amplitude of the differences between vaccines may be exaggerated. This is because there were different age-specific patterns in vaccine delivery (AstraZeneca having been primarily administered to older people) as well as differences in transmission dynamics for the age groups that received the different vaccines.

In conclusion, we found evidence of increasing prevalence of swab-positivity among school-aged children as well as higher prevalence of swab-positivity within three to six months following two-dose vaccination against COVID-19 in adults. Ongoing efforts to deliver single-dose vaccination to older school children, two doses to all adults, and booster doses to adults aged 50 years and over (as well as health and social care workers and younger people at risk) should help to counteract any reduction in immunity at both individual and population levels. The upcoming (typically) one-week half-term school holiday in England may have an impact on infection rates not just in children but their household members whose behaviours and social mixing patterns may be affected over the holiday period. In addition, we are entering the winter season in England when typically the NHS comes under strain from influenza and other infections. It is important that the vaccination programme maintains high coverage and reaches children and unvaccinated or partially vaccinated adults to reduce transmission and associated disruptions to work and education.

## Materials and Methods

The REACT study methods have been reported elsewhere [8]. Briefly, in REACT-1, we invite non-overlapping random cross-sectional samples of the population in England (aged 5 years and over) to take part, with data collection occurring monthly over a period of two to three weeks (except December 2020 and August 2021 when no survey was undertaken). At each round of data collection, named individuals from the NHS list of patients registered with a general practitioner in England are invited to take part, based on lists obtained from NHS Digital.

From beginning of May 2020 to beginning of May 2021 (round 1 to round 11) we aimed for approximately equal numbers of participants in each of 315 lower-tier local authorities (LTLAs) in England (combining the Isles of Scilly with Cornwall and the City of London with Westminster) but from round 12 (late May to early June 2021) onwards, we modified the sampling procedure to obtain a random sample in proportion to population at LTLA level. This increased the sampling in higher population density inner urban areas although prevalence reporting was unaffected as we re-weight the data at each round to be representative of England as a whole as described below.

We ask participants to provide a self-administered throat and nose swab (or parent/guardian obtained swab for children aged 5 to 12 years) following written and video instructions. Swabs are sent to a central laboratory for RT-PCR, and the test is considered positive either if both the two gene targets (N gene and E gene) used are detected or if N gene is detected with cycle threshold (Ct) value less than 37.

For round 14 we modified the way that the swab samples were handled. From round 1 to round 13 (June to July 2021) we used dry swabs which were refrigerated in the home and then sent chilled to the laboratory by courier for RT-PCR testing. In round 14, we switched to wet swabs in saline solution which were then either returned by post or via courier without the cold chain, although samples were refrigerated on arrival at the depot before being transported onward to the laboratory. Participants were allocated randomly on a 1:1 basis to either post or courier.

For people taking part in the study, we obtain age, sex, address and residential postcode from the NHS register and collect further information on demographics, health (including symptom reporting and contact with a known COVID-19 case) and lifestyle from an online or telephone questionnaire (available on the study website [16]). Participants are also asked for consent for linkage to their NHS records including data from the COVID-19 immunization programme.

Response rates are calculated as the percentage of those invited from whom we receive a valid swab result; this was 19.7% across all rounds, 11.7% for round 13 and 12.2% for round 14.

### Viral genome sequencing

Where there was sufficient sample volume and for N gene Ct values < 34, samples were sent frozen for viral genome sequencing at the Quadram Institute, Norwich, UK. The ARTIC protocol [17] was used for amplification of viral RNA and CoronaHiT for preparation of sequencing libraries [18]. The ARTIC bioinformatic pipeline [19] was used for analysis of sequencing data and lineages were assigned using PangoLEARN [20].

### Statistical Analyses

Statistical analyses were carried out in R [21]. We calculated unweighted (crude) prevalence by sociodemographic, occupational and other groups by dividing counts of swab-positivity (from RT-PCR) by the number of valid swabs returned in that group. We then use rim weighting [22] to obtain prevalence weighted to be representative of the population of England as a whole. Variables considered were: age (seven age groups), sex, deciles of the index of material deprivation (IMD), LTLA counts and ethnic group (nine categories: white; mixed / multiple ethnic groups; Indian; Pakistani; Bangladeshi; Chinese; any other Asian background; Black African / Caribbean / other; and any other ethnic group or missing). We used logistic regression to adjust for the potential confounding effects of covariates on prevalence estimates.

We used an exponential model of growth or decay to analyse trends in swab positivity over time, making the assumption that the number of positive samples (from the total number of samples) each day arose from a binomial distribution. The model takes the form *I*(*t*) = *I*_0_.*e*^*rt*^, where *I*(*t*) is the probability of swab positivity at time t, *I*_0_ is the probability of swab positivity on the first day of data collection per round and r is the growth rate. The binomial likelihood for *P* (out of *N*) positive tests on a given day is then *P* ∼*B* (*N,I*_0_.*e*^*rt*^,) based on day of swabbing or, if unavailable, day of sample collection (courier) or first scan of the sample by the Post Office if sent by post. We estimated posterior credible intervals using a bivariate No-U-Turn Sampler assuming uniform prior distributions on *I*_0_ and r [23]. We estimated the reproduction number R assuming a generation time that has a gamma distribution with shape parameter, n = 2.29 and rate parameter *β* = 0.36 (corresponding to a mean generation time of 6.29 days) [24]. We estimated R for round 14 from the equation *R* =(1+*r* / *β*)^^^ *n* [25] using data from all participants and stratified by age (5 to 17, 18 to 54, and 55+ years). We also estimated R for different definitions of swab-positivity and separately for samples sent by courier or post.

We fit a Bayesian penalised-spline (P-spline) model [26] to the daily data using a No U-Turns Sampler in logit space, with the data segmented into approximately 5 day sections by regularly spaced knots, and further knots included beyond the study period to minimise edge effects. We also fit P-splines to the REACT-1 data stratified by age as above in which a P-spline was fit separately to each age group but the smoothing parameter, ρ was assumed to be the best fitting value obtained for the model fit to all data.

We calculated VE against infection from combining data from round 13 and round 14 to increase statistical power. These estimates were based on data from linkage (with consent) to the national COVID-19 vaccination dataset which gave reliable dates of vaccination (unlike the self-report data where there were many missing dates). Based on the dates from data linkage, a person was considered to have had one dose 14 days after administration of the vaccine (before then being considered unvaccinated) and two doses 14 days after the second vaccination (before then being considered as having one dose only). We estimated VE as 1 - odds ratio, where the odds ratio was obtained from a logistic regression model comparing swab positivity among vaccinated and unvaccinated individuals, with adjustment for round, then sequentially round, age and sex, and round, age, sex, index of multiple deprivation quintile and ethnicity.

### Public involvement

A Public Advisory Panel provides input into the design, conduct and dissemination of the REACT research programme.

### Ethics

We obtained research ethics approval from the South Central-Berkshire B Research Ethics Committee (IRAS ID: 283787).

## Data Availability

Supporting data for tables and figures are available either: in this folder (https://drive.google.com/drive/folders/1jhukdN5_XPhoHcmL1HGpwCF1CWKhlhMa?usp=sharing); or in the following GitHub folder https://github.com/mrc-ide/reactidd/tree/master/inst/extdata/round_14_sup_date_figs

## Data availability

Supporting data for tables and figures are available either: in this folder; or in the following GitHub folder.

## Acknowledgements

MC-H and MW acknowledge support from the H2020-EXPANSE project (Horizon 2020 grant No 874627). MC-H and BB acknowledge support from Cancer Research UK, Population Research Committee Project grant ‘Mechanomics’ (grant No 22184 to MC-H). SR, CAD acknowledge support: MRC Centre for Global Infectious Disease Analysis, National Institute for Health Research (NIHR) Health Protection Research Unit (HPRU), Wellcome Trust (200861/Z/16/Z, 200187/Z/15/Z), and Centres for Disease Control and Prevention (US, U01CK0005-01-02). GC is supported by an NIHR Professorship. HW acknowledges support from an NIHR Senior Investigator Award and the Wellcome Trust (205456/Z/16/Z). PE is Director of the Medical Research Council (MRC) Centre for Environment and Health (MR/L01341X/1, MR/S019669/1). PE acknowledges support from Health Data Research UK (HDR UK); the NIHR Imperial Biomedical Research Centre; NIHR HPRUs in Chemical and Radiation Threats and Hazards, and Environmental Exposures and Health; the British Heart Foundation Centre for Research Excellence at Imperial College London (RE/18/4/34215); and the UK Dementia Research Institute at Imperial College London (MC_PC_17114). We thank The Huo Family Foundation for their support of our work on COVID-19.

We thank key collaborators on this work – Ipsos MORI: Kelly Beaver, Sam Clemens, Gary Welch, Nicholas Gilby, Kelly Ward, Galini Pantelidou and Kevin Pickering; Institute of Global Health Innovation at Imperial College London: Gianluca Fontana, Sutha Satkunarajah, Didi Thompson and Lenny Naar; School of Public Health, Imperial College London: Eric Johnson, Rob Elliott, Graham Blakoe; North West London Pathology and Public Health England for help in calibration of the laboratory analyses; Patient Experience Research Centre at Imperial College London and the REACT Public Advisory Panel; Quadram Institute, Norwich, UK: Thanh Le Viet, Nabil-Fareed Alikhan, Leigh M Jackson, Catherine Ludden; NHS Digital for access to the NHS register; the Department of Health and Social Care for logistic support; and the COVID-19 Taskforce of the Royal Statistical Society (UK) for helpful comments. SR acknowledges helpful discussion with attendees of meetings of the UK Government Scientific Pandemic Influenza – Modelling (SPI-M) committee.

Quadram authors gratefully acknowledge the support of the Biotechnology and Biological Sciences Research Council (BBSRC); their research was funded by the BBSRC Institute Strategic Programme Microbes in the Food Chain BB/R012504/1 and its constituent project BBS/E/F/000PR10352. We thank members of the COVID-19 Genomics Consortium UK (COG-UK) for their contributions to generating the genomic data used in this study. COG-UK is supported by funding from the MRC, part of UK Research & Innovation (UKRI), NIHR and Genome Research Limited, operating as the Wellcome Sanger Institute.

## Additional information

Full list of COG-UK author’s names and affiliations are available [here](https://docs.google.com/document/d/13kv6XftpEba0RW6xu5xCLeIlsmbzp5I4/edit?usp=sharing&ouid=110640444141243456444&rtpof=true&sd=true)

## Supplementary Materials

**Table S1.** Unweighted and weighted prevalence of swab-positivity and Median N-gene Ct values between individuals with different methods of swab test collection in round 14.

| Data | Negatives | Positives | Total | Unweighted prevalence |  | Weighted prevalence |  | Median N-gene Ct value* | P-value** |
| --- | --- | --- | --- | --- | --- | --- | --- | --- | --- |
| Post | 53428 | 394 | 53822 | 0.73% | ( 0.66% , 0.81% ) | 0.80% | ( 0.72% , 0.90% ) | 28.9 ( 27.9 , 29.9 ) | ref |
| Courier | 46335 | 370 | 46705 | 0.79% | ( 0.71% , 0.88% ) | 0.85% | ( 0.76% , 0.96% ) | 28.0 ( 27.0 , 29.0 ) | 0.06 |

\* 95% Confidence intervals in the median were estimated using quantile regression
\*\*P-values were calculated using the two-sided Wilcoxon (also known as Mann Whitney) test and are relative to the distribution of Ct values in individuals that sent their test via post

**Table S2.** Proportion of each Delta sub-lineage detected in 475 (62.1%) positive samples from round 14^*^.

| Sub-lineage | (Total 475) | Proportion |
| --- | --- | --- |
| B.1.617.2 (Delta/Sub-lineage not known) | 129 | 0.272 ( 0.234 , 0.313 ) |
| AY.26 | 13 | 0.027 ( 0.016 , 0.046 ) |
| AY.3 | 1 | 0.002 ( 0.000 , 0.012 ) |
| AY.4 | 292 | 0.615 ( 0.570 , 0.657 ) |
| AY.4.1 | 7 | 0.015 ( 0.007 , 0.030 ) |
| AY.5 | 19 | 0.040 ( 0.026 , 0.062 ) |
| AY.6 | 8 | 0.017 ( 0.009 , 0.033 ) |
| AY.9 | 2 | 0.004 ( 0.001 , 0.015 ) |
| AY.33 | 1 | 0.002 ( 0.000 , 0.012 ) |
| AY.36 | 1 | 0.002 ( 0.000 , 0.012 ) |
| AY.11 | 1 | 0.002 ( 0.000 , 0.012 ) |
| AY.20 | 1 | 0.002 ( 0.000 , 0.012 ) |

\* One Delta variant with E484K mutation detected

**Figure S1.**
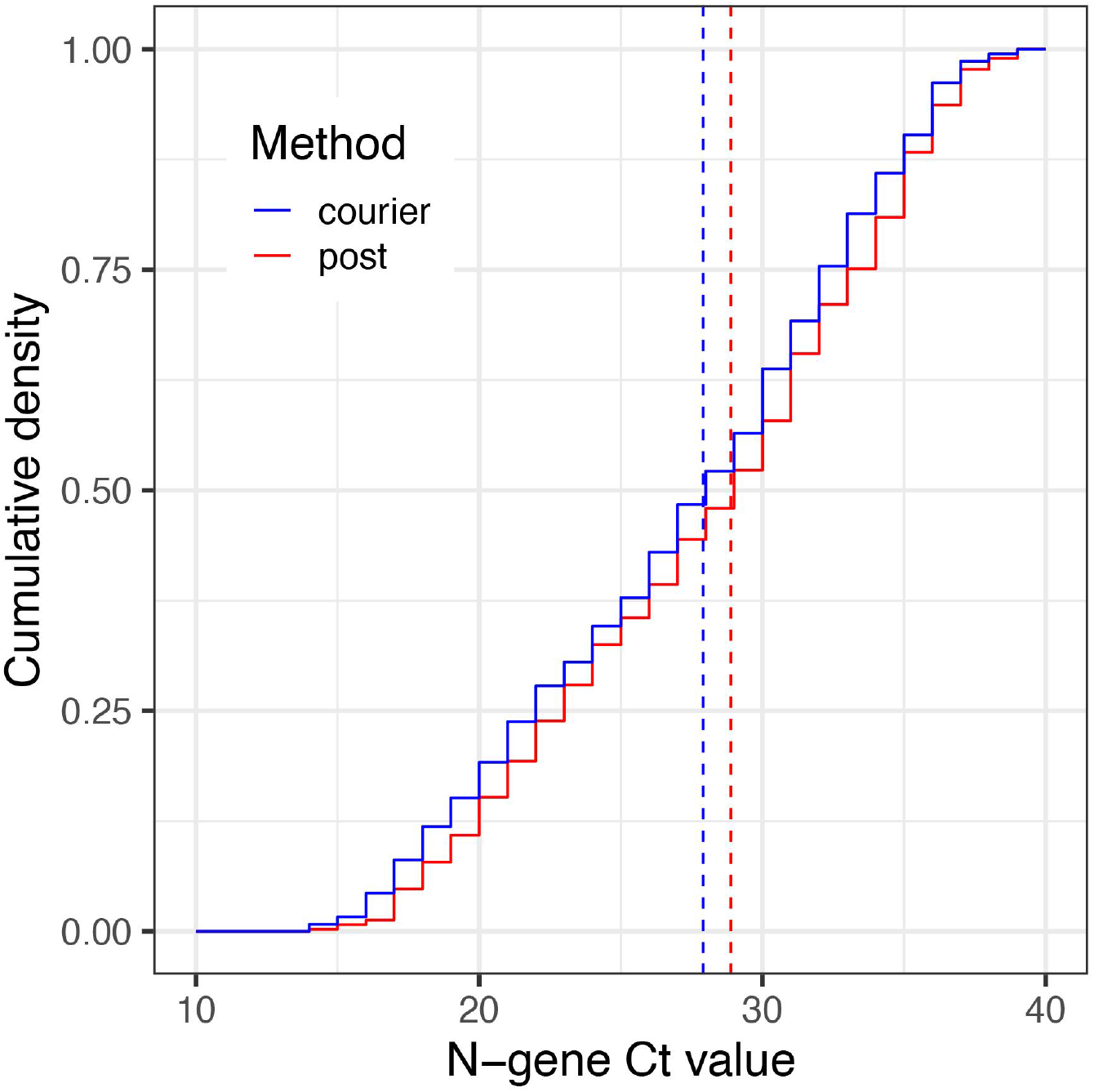
Distribution of N-gene Ct values, by method used in collecting swab tests from participants, for all positive samples. Cumulative distribution of all N-gene Ct values for those whose swab test was collected by courier (blue) and those who sent their swab test in the post (red).

## References

1. COVID-19 map - Johns Hopkins Coronavirus resource Center. [cited 13 Oct 2021]. Available: https://coronavirus.jhu.edu/map.html

2. Riley S, Wang H, Eales O, Haw D, Walters CE, Ainslie KEC, et al. REACT-1 round 12 report: resurgence of SARS-CoV-2 infections in England associated with increased frequency of the Delta variant. bioRxiv. medRxiv; 2021. doi:10.1101/2021.06.17.21259103

3. UK Government. COVID-19 Data Dashboard (Cases). [cited 13 Oct 2021]. Available: https://coronavirus.data.gov.uk/details/cases

4. UK Government. COVID-19 Data Dashboard (vaccination data). [cited 13 Oct 2021]. Available: https://coronavirus.data.gov.uk/details/vaccinations

5. Schools COVID-19 operational guidance. [cited 13 Oct 2021]. Available: https://www.gov.uk/government/publications/actions-for-schools-during-the-coronavirus-outbreak/schools-covid-19-operational-guidance

6. BBC News. First Scottish pupils back at school for new term beyond level zero. BBC. 11 Aug 2021. Available: https://www.bbc.co.uk/news/uk-scotland-58163742. Accessed 13 Oct 2021.

7. Riley S, Ainslie KEC, Eales O, Walters CE, Wang H, Atchison C, et al. Resurgence of SARS-CoV-2: Detection by community viral surveillance. Science. 2021;372: 990–995.

8. Riley S, Atchison C, Ashby D, Donnelly CA, Barclay W, Cooke G, et al. REal-time Assessment of Community Transmission (REACT) of SARS-CoV-2 virus: Study protocol. Wellcome Open Research. 2020. p. 200. doi:10.12688/wellcomeopenres.16228.1

9. Latest insights team. Coronavirus (COVID-19) latest insights - Office for National Statistics. Office for National Statistics; 7 Oct 2021 [cited 13 Oct 2021]. Available: https://www.ons.gov.uk/peoplepopulationandcommunity/healthandsocialcare/conditionsanddiseases/articles/coronaviruscovid19latestinsights/infections

10. BBC News. Covid: People 45 or over in England invited to book vaccine. BBC. 13 Apr 2021. Available: https://www.bbc.co.uk/news/uk-56729897. Accessed 13 Oct 2021.

11. Lopez Bernal J, Andrews N, Gower C, Gallagher E, Simmons R, Thelwall S, et al. Effectiveness of Covid-19 Vaccines against the B.1.617.2 (Delta) Variant. N Engl J Med. 2021. doi:10.1056/NEJMoa2108891

12. Elliott P, Haw D, Wang H, Eales O, Walters CE, Ainslie KEC, et al. REACT-1 round 13 final report: exponential growth, high prevalence of SARS-CoV-2 and vaccine effectiveness associated with Delta variant in England during May to July 2021. bioRxiv. 2021. doi:10.1101/2021.09.02.21262979

13. Public Health England. COVID-19 vaccine surveillance reports (weeks 19 to 38). http://GOV.UK; 13 May 2021 [cited 13 Oct 2021]. Available: https://www.gov.uk/government/publications/covid-19-vaccine-surveillance-report

14. Bar-On YM, Goldberg Y, Mandel M, Bodenheimer O, Freedman L, Kalkstein N, et al. Protection of BNT162b2 Vaccine Booster against Covid-19 in Israel. N Engl J Med. 2021;385: 1393–1400.

15. COVID-19 vaccination: a guide to booster vaccination. [cited 13 Oct 2021]. Available: https://www.gov.uk/government/publications/covid-19-vaccination-booster-dose-resources/covid-19-vaccination-a-guide-to-booster-vaccination

16. REal-time Assessment of Community Transmission (REACT) study. In: Imperial College [Internet]. Available: https://www.imperial.ac.uk/medicine/research-and-impact/groups/react-study/

17. Quick J. nCoV-2019 sequencing protocol v3 (LoCost). 2020 [cited 4 May 2021]. Available: https://www.protocols.io/view/ncov-2019-sequencing-protocol-v3-locost-bh42j8ye

18. Baker DJ, Aydin A, Le-Viet T, Kay GL, Rudder S, de Oliveira Martins L, et al. CoronaHiT: high-throughput sequencing of SARS-CoV-2 genomes. Genome Med. 2021;13: 21.

19. A Nextflow pipeline for running the ARTIC network’s field bioinformatics tools. Github; Available: https://github.com/connor-lab/ncov2019-artic-nf

20. Phylogenetic Assignment of Named Global Outbreak LINeages (PANGOLIN). Github; Available: https://github.com/cov-lineages/pangolin

21. R Core Team. R: A Language and Environment for Statistical Computing. R Foundation for Statistical Computing; 2020. Available: https://www.R-project.org/

22. Sharot T. Weighting survey results. 1986 [cited 12 Jan 2021]. Available: http://redresearch.com/wp/wp-content/uploads/2016/01/Weighting-Survey-Results.pdf

23. Hoffman MD, Gelman A. The No-U-Turn Sampler: Adaptively Setting Path Lengths in Hamiltonian Monte Carlo. arXiv [stat.CO]. 2011. Available: http://arxiv.org/abs/1111.4246

24. Bi Q, Wu Y, Mei S, Ye C, Zou X, Zhang Z, et al. Epidemiology and transmission of COVID-19 in 391 cases and 1286 of their close contacts in Shenzhen, China: a retrospective cohort study. Lancet Infect Dis. 2020;20: 911–919.

25. Wallinga J, Lipsitch M. How generation intervals shape the relationship between growth rates and reproductive numbers. Proceedings Of The Royal Society B-Biological Sciences. 2006;274: 599–604.

26. Lang S, Brezger A. Bayesian P-Splines. J Comput Graph Stat. 2004;13: 183–212.

